# Acute Social Stress Potentiates Cue-Driven Behavior in Alcohol Use Disorder: Behavioral and Neural Evidence

**DOI:** 10.64898/2026.09.01.26361915

**Authors:** Maren Born, Samanda Krasniqi, Carlotta Riemerschmid, Hao Chen, Louis Thill, Michael N. Smolka, Florian Schlagenhauf, Andreas Heinz, Eva Friedel, Claudia Ebrahimi, Maria Garbusow

## Abstract

**Background:** Pavlovian-to-instrumental transfer (PIT) describes how conditioned cues shape instrumentally learned behavior and has been linked to cue-driven alcohol seeking and relapse in alcohol use disorder (AUD). Acute psychosocial stress is an established relapse risk factor that may amplify cue-driven motivational control. Whether acute stress modulates PIT behaviorally and neurally, and whether this differs in AUD, remains unclear.

**Methods:** Within the TRR265 study, 109 participants (62 AUD, 46 controls (CON)) completed a within-subject, randomized, two-day PIT paradigm under Trier Social Stress Test and placebo conditions, with repeated salivary cortisol sampling across sessions. A subsample (n = 62; 34 AUD, 28 CON) underwent fMRI during the transfer phase.

**Results:** A robust PIT effect emerged across groups, and acute stress significantly enhanced cue-driven response vigor (p = .034), with no AUD-specific behavioral effect. Exploratorily, cortisol responders showed stronger stress-related PIT enhancement than non-responders (p = .022). Neurally, PIT engaged the right amygdala during placebo. Acute stress induced less downregulation of PIT-related left amygdala activity in AUD than in CON (p_FWE_ = .019), with no group difference during placebo.

**Conclusions:** Acute psychosocial stress potentiated the motivational influence of non-drug-related conditioned cues on behavior irrespective of AUD status, identifying PIT as a candidate mechanism through which stress may promote alcohol-seeking behavior. Despite comparable behavioral effects, AUD was associated with blunted stress-induced downregulation of PIT-related amygdala activity, suggesting altered neural regulation of cue-driven processing whose relevance for future alcohol consumption and clinical outcome remains to be established.

## INTRODUCTION

Individuals with alcohol use disorder (AUD) repeatedly encounter environmental cues that can acquire motivational significance, subsequently trigger craving and promote substance-seeking behavior despite intentions to reduce or abstain from consumption [1–3]. It is therefore critical to reveal the mechanisms through which learned cues acquire motivational control over behavior, both to explain difficulties to reduce drinking and remain abstinent and to inform the development of targeted interventions for AUD behavior [4]. Incentive-sensitization theory proposes that repeated drug exposure can attribute excessive incentive salience on reward-predictive cues, allowing them to capture attention and trigger motivational responses independently of their hedonic value [5, 6]. Pavlovian-to-instrumental transfer (PIT) provides an experimental framework for quantifying how conditioned cues influence independently learned instrumental behavior [7, 8]. Whereas drug-related PIT probes the influence of drug-associated cues on behavior, non-drug-related PIT assesses whether cue-driven motivational processes generalize beyond substance-specific stimuli. As such, it captures a more general mechanism of motivational control that is not contingent on alcohol-related cues or craving and may therefore provide a translational measure.

Converging evidence suggests that non-drug PIT captures motivational processes relevant to AUD and its clinical course [4, 9]. In humans, stronger non-drug-related PIT has been demonstrated in detoxified individuals with AUD compared with controls (CON) [10], as well as in high- versus low-risk drinkers, with PIT strength additionally associated with polygenic risk for alcohol consumption [11]. Importantly, PIT-related Nucleus accumbens (NAcc) activation prospectively predicted relapse and subsequent alcohol intake in individuals with AUD [12, 13]. Moreover, stronger alcohol approach bias was associated with both heightened behavioral PIT and greater PIT-related NAcc activity [14].

A critical unresolved question however is how this mechanism operates under stress. Psychosocial stress is a well-established determinant of AUD trajectories and relapse risk (Brown et al., 1990; Sinha, 2008, 2012; Sinha et al., 2011), and conditioned cues are frequently encountered in contexts of heightened stress. Rather than constituting independent risk processes, it has been hypothesized that stress may alter the extent to which conditioned cues exert motivational control over behavior [4, 15]. This interaction is also neurobiologically plausible: stress engages corticolimbic and mesolimbic circuits that overlap with neural systems underlying PIT, including the amygdala and ventral striatum [16]. In parallel, acute stress can shift behavioral control away from flexible, goal-directed actions toward more automatic responses [17, 18].

Experimental evidence for such stress-dependent modulation of PIT remains limited and inconsistent. In rodents, intra-accumbens administration of corticotropin-releasing factor (CRF), mimicking the neuroendocrine stress response, increased PIT for sucrose reward by magnifying the incentive salience of Pavlovian cues [19]. In humans, acute stress selectively increased cue-triggered “wanting” independently of subjective “liking” [20], while stress and anxiety have also been associated with altered Pavlovian control over instrumental behavior [21]. Conversely, other studies report no such effects. Chronic unpredictable stress transiently impaired, rather than enhanced, PIT in rats [22], while also acute stressors did not alter the PIT effect in rats [23]. Pritchard et al. found that negative emotional appraisal left PIT itself unaffected, but instead disrupted outcome devaluation effects on instrumental choice [24].

Critically, the only previous study directly examining interaction in a substance use disorder (SUD) found no modulation through stress of smoking-related PIT in smokers [25]. Thus, whether acute psychosocial stress potentiates cue-driven motivational control in AUD, and how this is reflected in the underlying neural circuits, remains unclear.

The present study investigated the effects of acute psychosocial stress on non-drug-related PIT in individuals with AUD and matched CON, combining behavioral, endocrine, and functional neuroimaging measures in a within-subject stress versus no-stress design. We used a paradigm that yields two complementary behavioral expressions of PIT. Motivational PIT captures the extent to which Pavlovian cues invigorate instrumental responding, reflecting cue-driven motivational processes. In contrast, interference PIT quantifies the extent to which Pavlovian cues conflict with the required instrumental responses, indexing the increase in error rates on incongruent relative to congruent trials [26]. Although behaviorally correlated, previous work indicates that two measures rely on partially distinct neural mechanisms, with motivational PIT primarily engaging the amygdala and nucleus accumbens (NAcc), whereas interference PIT additionally recruits cognitive-control networks [26, 27]. Because stress has been proposed to amplify the motivational impact of reward-predictive cues [20], our hypotheses focused on motivational PIT. We expected stress to enhance motivational PIT, particularly in AUD. Interference PIT was included as an complimentary and explorative outcome to determine whether stress influenced Pavlovian-instrumental conflict in addition to motivational invigoration and is reported in the Supplements.

## METHODS

### Participants

This study was conducted as part of the TRR 265 consortium (https://www.trr265.org/sfb-trr-265, ClinicalTrials.gov ID: NCT05992272) [28]. Data collection was carried out in Berlin, Germany, with approval from local ethics committees of the Charité Universitätsmedizin Berlin (EA2/239/18). We included a total of 109 participants with complete behavioral data, comprising 62 individuals with AUD and 46 CON, matched for age and gender. Following standard fMRI quality control procedures (see Supplements), the final neuroimaging sample consisted of 62 participants (34 AUD, 28 CON). Each participant completed two experimental sessions (stress, no-stress) on consecutive days in a randomized order. For detailed inclusion and exclusion criteria, see Supplements. Demographics and clinical characteristics are shown in Table 1 and in the Supplements.

**Table 1.**
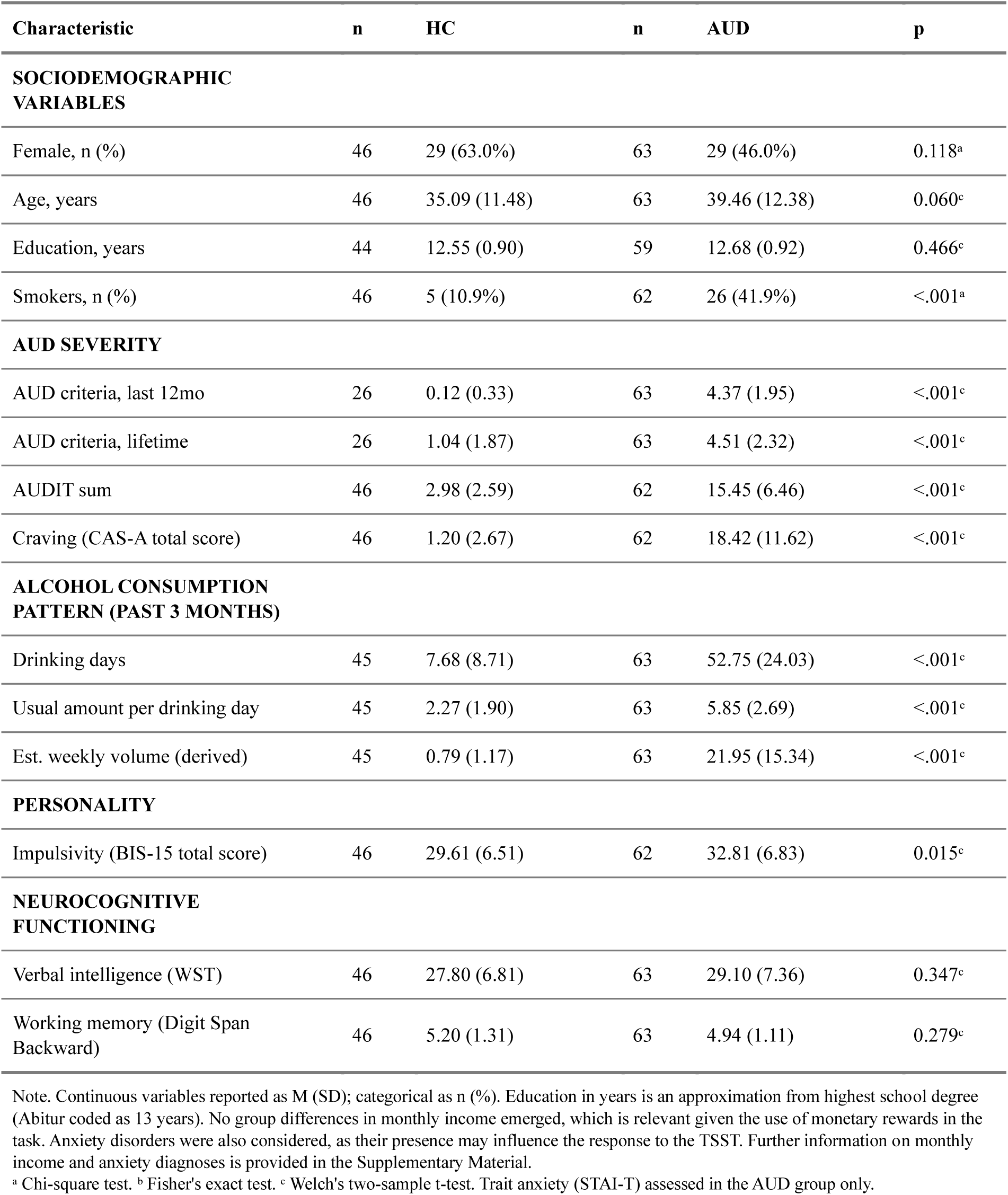
Sample Characteristics.

| Characteristic | n | HC | n | AUD | p |
| --- | --- | --- | --- | --- | --- |
| **SOCIODEMOGRAPHIC VARIABLES** |  |  |  |  |  |
| Female, n (%) | 46 | 29 (63.0%) | 63 | 29 (46.0%) | 0.118<sup>a</sup> |
| Age, years | 46 | 35.09 (11.48) | 63 | 39.46 (12.38) | 0.060<sup>c</sup> |
| Education, years | 44 | 12.55 (0.90) | 59 | 12.68 (0.92) | 0.466<sup>c</sup> |
| Smokers, n (%) | 46 | 5 (10.9%) | 62 | 26 (41.9%) | <.001<sup>a</sup> |
| **AUD SEVERITY** |  |  |  |  |  |
| AUD criteria, last 12mo | 26 | 0.12 (0.33) | 63 | 4.37 (1.95) | <.001<sup>c</sup> |
| AUD criteria, lifetime | 26 | 1.04 (1.87) | 63 | 4.51 (2.32) | <.001<sup>c</sup> |
| AUDIT sum | 46 | 2.98 (2.59) | 62 | 15.45 (6.46) | <.001<sup>c</sup> |
| Craving (CAS-A total score) | 46 | 1.20 (2.67) | 62 | 18.42 (11.62) | <.001<sup>c</sup> |
| **ALCOHOL CONSUMPTION PATTERN (PAST 3 MONTHS)** |  |  |  |  |  |
| Drinking days | 45 | 7.68 (8.71) | 63 | 52.75 (24.03) | <.001<sup>c</sup> |
| Usual amount per drinking day | 45 | 2.27 (1.90) | 63 | 5.85 (2.69) | <.001<sup>c</sup> |
| Est. weekly volume (derived) | 45 | 0.79 (1.17) | 63 | 21.95 (15.34) | <.001<sup>c</sup> |
| **PERSONALITY** |  |  |  |  |  |
| Impulsivity (BIS-15 total score) | 46 | 29.61 (6.51) | 62 | 32.81 (6.83) | 0.015<sup>c</sup> |
| **NEUROCOGNITIVE FUNCTIONING** |  |  |  |  |  |
| Verbal intelligence (WST) | 46 | 27.80 (6.81) | 63 | 29.10 (7.36) | 0.347<sup>c</sup> |
| Working memory (Digit Span Backward) | 46 | 5.20 (1.31) | 63 | 4.94 (1.11) | 0.279<sup>c</sup> |

Note. Continuous variables reported as M (SD); categorical as n (%). Education in years is an approximation from highest school degree (Abitur coded as 13 years). No group differences in monthly income emerged, which is relevant given the use of monetary rewards in the task. Anxiety disorders were also considered, as their presence may influence the response to the TSST. Further information on monthly income and anxiety diagnoses is provided in the Supplementary Material.
<sup>a</sup> Chi-square test. <sup>b</sup> Fisher's exact test. <sup>c</sup> Welch's two-sample t-test. Trait anxiety (STAI-T) assessed in the AUD group only.

#### PIT Paradigm

The PIT paradigm was based on a design previously established by our group [29]. See Supplementary Material for details and Figure 1 for an overview.

**Figure 1.**
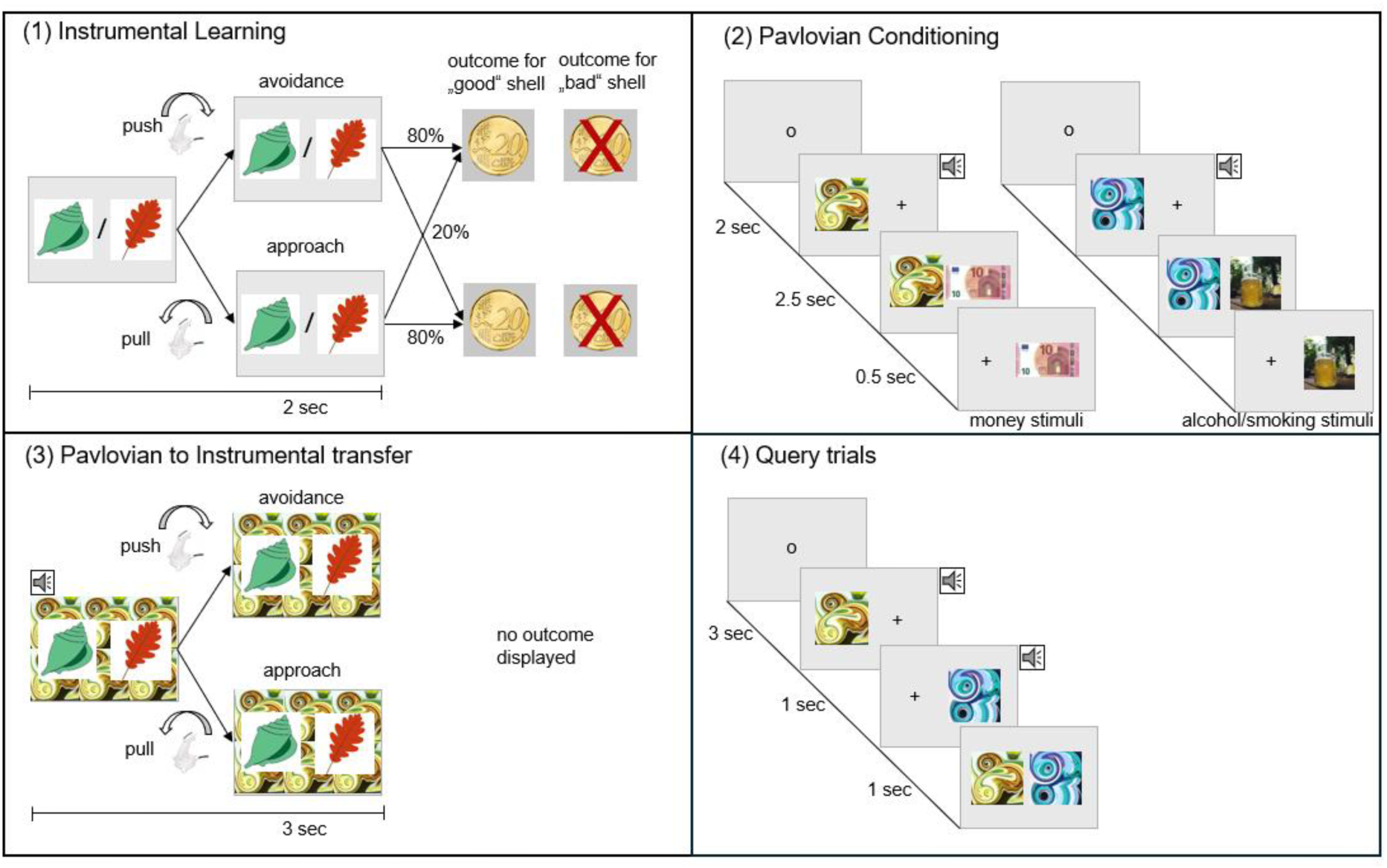
Task design. The PIT task comprised four phases: **(1) Instrumental training:** participants learned to collect “good” shells or leafs (approach trials) and reject “bad” ones (avoid trials) via immediate probabilistic feedback; **(2) Pavlovian Conditioning:** abstract fractals paired with auditory tones (CS) were associated with monetary outcomes (appetitive: +10€, neutral: 0€, aversive: −10€) or drug-related images (alcohol, smoking) as unconditioned stimuli (US); **(3) PIT:** across 180 trials participants performed the instrumental task under nominal extinction while Pavlovian CS were presented in the background; **(4) Query trials:** to assess implicit contingency knowledge, participants had to choose between two CSs based upon their liking. The task was programmed in MATLAB using the Psychophysics Toolbox and presented on a laptop. Participants responded with their right hand using an MRI-compatible joystick

### MRI Acquisition

Structural and functional MRI data were acquired on 3-Tesla Siemens Trio scanners (Siemens AG, Erlangen, Germany). Details can be found in the Supplements.

### Acute Social Stress Intervention

The Trier Social Stress Test (TSST), a widely recognized protocol for eliciting acute social stress, was implemented with modifications to meet COVID-19 safety guidelines that were in effect during data acquisition [30–32]. On the alternate experimental day, participants underwent a placebo version, designed to mimic the structure of the TSST without inducing stress [33]. The order of intervention was randomized across participants.

### Salivary Cortisol

To assess the responsiveness of the hypothalamic-pituitary-adrenal (HPA) axis to acute social stress, salivary cortisol samples were collected at seven timepoints on each experimental day (see Figure 2). The samples were frozen and stored at -20°C in Salivette® collection devices (Sarstedt, Nümbrecht, Germany) and centrifuged for five minutes at 3,000 rpm to create a clear supernatant with low viscosity. Salivary cortisol concentrations were analyzed using a chemiluminescence immunoassay with high sensitivity (IBL International, Hamburg, Germany) and within-run coefficients of variation ranging from 3.0% to 5.0% and between-run coefficients of variation ranging from 2.9% to 5.0%.

**Figure 2.**
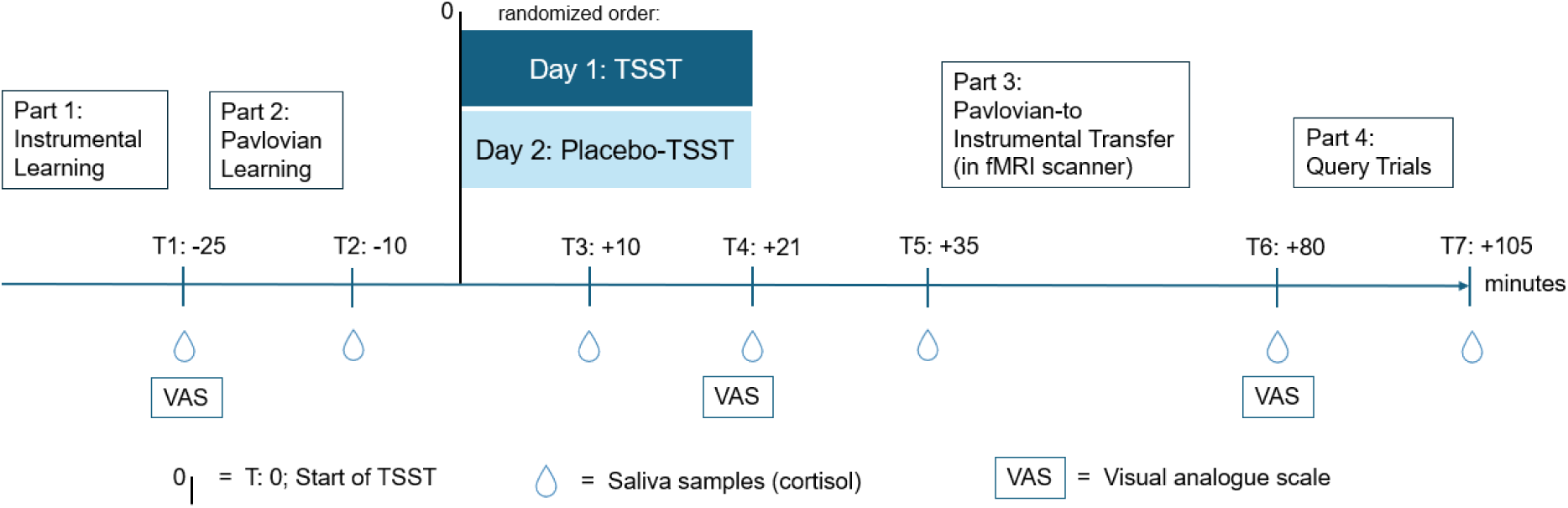
Two-day study design. Based on prior evidence, peak stress effects were expected approximately 10 minutes following TSST completion [33]. **S**aliva samples were collected at seven time points relative to the start of the (Placebo-)TSST at T= 0 minutes (min): T1 (−21 min; between Instrumental Learning and Pavlovian Conditioning), T2 (−10 min; after Pavlovian Conditioning), T3 (+10 min; immediately before the free speech task of the TSST), T4 (+20 min; immediately after the TSST), T5 (+35 min; upon arrival at Berlin Center of Advanced Neuroimaging (BCAN)), T6 (+80 min; after PIT and fMRI imaging), and T7 (+105 min; following the debriefing questionnaire).

**Figure 3.**
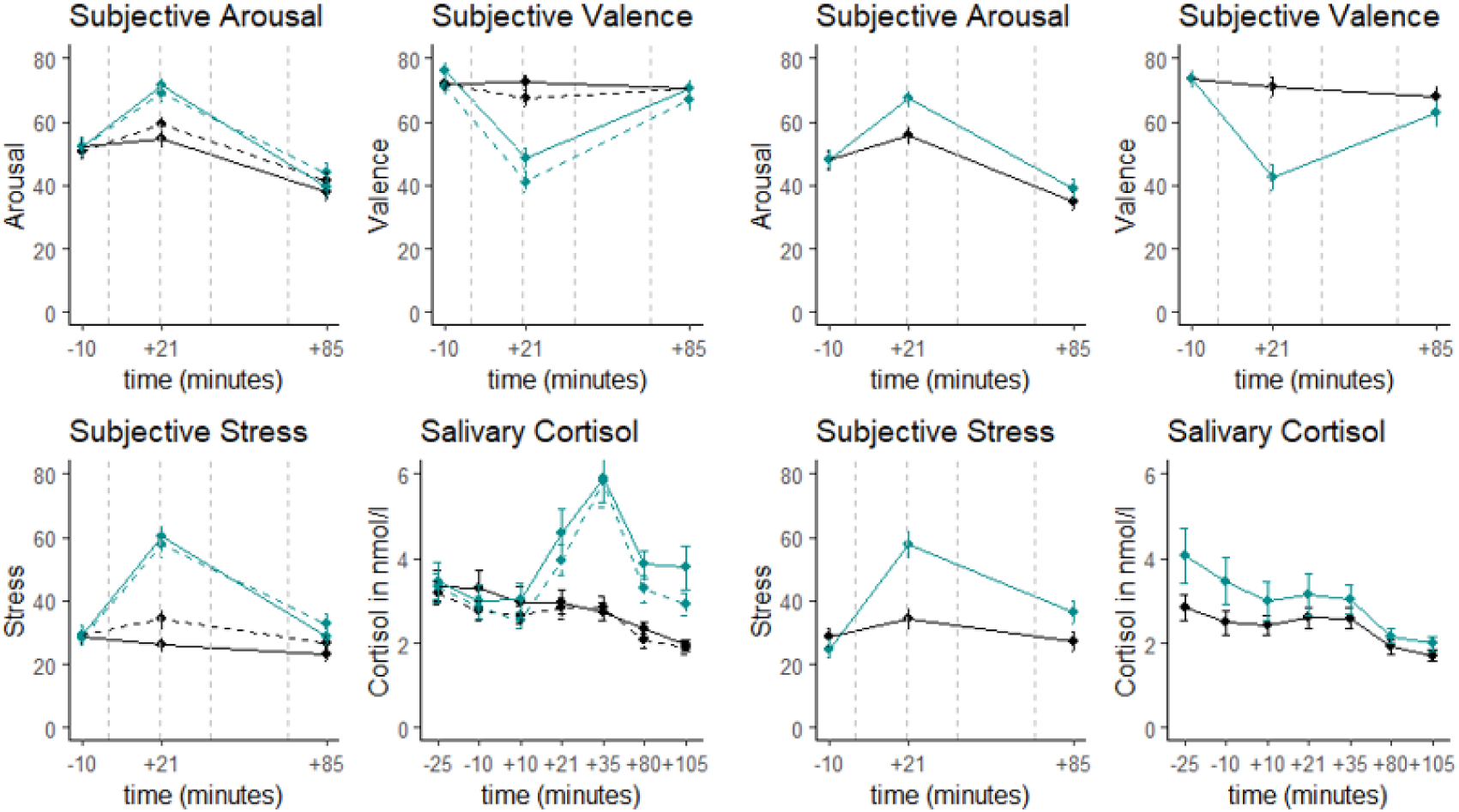
Physiological and Subjective Stress Responses in the Full Sample (left 3.1) and the Cortisol Non-Responder Subgroup (right 3.2). Figure 3.1.: Dashed Line = Controls (CON) , Solid Line = participants with AUD, Figure 3.1. and 3.2: Cyan Green= TSST, Black = Placebo-TSST

### Subjective Stress Response

On both experimental days participants were asked at three timepoints (T2 (-10), T4 (+21), T6 (+80), Figure 2) to complete a visual analogue scale (VAS) measuring subjective arousal (“do you feel active or sleepy?”), valence (“do you feel happy or unhappy?”) and stress (“do you feel stressed or not stressed ?”) on a range from 0 (sleepy, unhappy, or not stressed) to 100 (active, happy, or stressed).

### Data Analysis

Statistical analyses were performed in MATLAB R2022b and R (v4.4.1, RStudio v2024.02.04). For fMRI data the Statistical Parametric Mapping software package (SPM12) was used.

### Salivary Cortisol Analysis

For cortisol response analysis we calculated the area under the curve with respect to ground (AUCg) for the seven samples using the standard trapezoid formula [34]. Repeated-measures ANOVAs assessed effects of intervention (stress, no-stress: within-subject) and group (AUD, CON: between-subject) on AUCg and baseline cortisol levels, as defined as the first sample [35]. Consistent with known interindividual variability in cortisol response to acute social stress [36, 37] and following established methods [38], we classified participants who showed a cortisol increase of ≥1.5 nmol/L post-TSST (T4–T7) relative to the lowest pre-TSST level (T1–T2) as responders. Subsequently, responder status was included in exploratory behavioral analysis. For missing data handling, see Supplements.

### Subjective Ratings

To quantify changes in all three measures (arousal, valence, stress), peak values were calculated by subtracting the pre-intervention values (T2 (-10)) from the post-intervention values (T4 (+21)). Subsequently, peak arousal, peak valence, and peak stress were separately used as dependent variables in repeated-measures ANOVAs with intervention (TSST, Placebo-TSST) as the within-group factor and group (AUD, CON) as the between-group factor.

### Behavioral Analysis

While the primary focus of behavioral analysis was on PIT, we analyzed Instrumental and Pavlovian Learning to ensure comparable learning effects across groups. (Supplements X). For motivational PIT, linear mixed-effects models, implemented in the lme4 package, were conducted for monetary Pavlovian CS trials [39]. The drug-related trials (alcohol, smoking) will be analyzed in future work. Peak velocity of joystick movement, a measure of instrumental response vigor, was predicted by the contrast-coded fixed effects instrumental condition (approach = 0.5, avoid = -0.5), intervention (TSST = 0.5, Placebo = -0.5) and group (AUD = 0.5, CON = -0.5 or responder = 0.5, non-responder -0.5). Pavlovian CS was treated as a numeric variable (-1 for -10€, 0 for neutral, +1 for +10€). For the optimal random effect structure, we systematically compared models of increasing complexity using likelihood ratio tests [40, 41]. The final model incorporated random intercepts for subject, day nested within subject, Pavlovian fractal (fractal type) and instrumental stimuli (leaf/shell type) to control for item specific differences, alongside random slopes for Pavlovian CS and instrumental condition within subject. PIT regression slopes were extracted for each day individually. For further details and PIT Interference results, see Supplements.

### Imaging Analysis

Following established preprocessing procedures (see Supplements), individual first-level general linear models (GLMs) were implemented in SPM12 using an event-related design for non-drug related motivational PIT. Both experimental sessions were modelled within a single design matrix to capture within-subject variability and ensure consistent estimation of task regressors, yielding one beta estimate per regressor and participant.

Two main task regressors represented instrumental approach and avoid trials, each with three serially entered parametric modulators: (1) the PIT parameter (Pavlovian CS value × peak velocity), capturing the trial-by-trial PIT effect; (2) Pavlovian CS value and (3) peak velocity. Onsets were defined by PIT trial start. Drug-related trials were modelled in a separate regressor of no interest. To account for motor-related variance, peak velocity was included as an additional regressor with beginning of movement as onset. Trials with missing responses or outlier trials were modelled separately. All regressors were handled as stick-function convolved with the canonical HRF. First-level models further included 24 head motion parameters (six realignment regressors, their derivatives and squared terms).

At the second level, one-sample and two-sample t-tests were conducted to assess the PIT effect and group differences (AUD vs. CON). Contrasts were set on approach and avoid trials simultaneously to capture the PIT effect across all non-drug related trials. Intervention effects (Stress > No-stress) were modelled within-subject using weighted contrasts (+1, -1) and assessed via two-tailed t-tests. Following the approach from previous work [11], we employed a region-of-interest (ROI) analyses that focused on bilateral Amygdala and NAcc. Both regions were selected a priori based on their established roles in Pavlovian-to-Instrumental-transfer [7, 42–45].

As done as in preceding PIT motivational publications, bilateral masks were generated using the Wake Forest University PickAtlas toolbox (https://www.nitrc.org/projects/wfu_pickatlas/, accessed April 30, 2025) [11]. Within the two ROIs, statistical inference was performed using small-volume correction (SVC; p < .05, FWE-corrected). Additionally, mean parametric estimates for the PIT parametric modulator were extracted from the bilateral Amygdala and the bilateral NAcc ROI for the no-stress as well as for the stress>no-stress contrast and correlated with individual behavioral PIT slopes, to examine brain-behavior relationship.

Exploratory whole brain analysis were performed applying an uncorrected voxel-level threshold of p < .001 with a minimum cluster extent of k ≥ 30 contiguous voxels (see Supplements).

## RESULTS

### Salivary Cortisol Response

Due to a Shapiro-Wilk test that indicated non-normality (W = 0.702, p < 0.001), cortisol values (nmol/L) were log transformed at subject level (N= 105). The repeated-measures ANOVA on AUCg revealed a robust main effect of intervention, with elevated salivary cortisol levels following the TSST (F(1, 103) = 51.29, p < .001, η² = .11). Contrary to hypotheses, there was no main effect of group (F(1, 103) = 0.08, p = .783, η² < .001) and no group × intervention interaction (F(1, 103) = 0.22, p = .642, η² < .001). Likewise, baseline cortisol levels did not differ between groups (F(1, 103) = 0.59, p = .443, η² < .001). Post-TSST, 64.76% of participants showed a cortisol increase ≥1.5 nmol/L (CON: 55.55%; AUD: 71.66%).

### Subjective Ratings

Stress induced significant changes across all subjective measures, with significant intervention effects for peak arousal (F(1, 104) = 32.87, p < .001, η²ₚ = .13), peak stress (F(1, 104) = 100.72, p < .001, η²ₚ = .34), and peak valence (F(1, 104) = 82.03, p < .001, η²ₚ = .30).

No significant main effects of group or group × intervention interactions were found for any subjective outcome. In the cortisol non-responder subgroup, an exploratory repeated-measures ANOVA confirmed a significant increase in perceived stress following the TSST (F(1, 36) = 29.85, p < .001, η²ₚ = .45). Importantly, subjective responses to the TSST did not significantly differ between cortisol responders and non-responders (see Supplementary Results).

### Behavioral Analysis 118

A robust motivational PIT effect emerged in monetary trials across both groups with higher Pavlovian stimulus values associated with increased joystick peak velocity (estimate = 211.70, *t* = 5.22, *p* < .001; Figure 4). Most notably, acute psychosocial stress significantly amplified this effect, as evidenced by a Pavlovian stimulus value × stress condition interaction (estimate = 53.36, *t* = 2.13, *p* = .034), indicating enhanced cue-driven invigoration of behavior under stress. No significant interactions involving group were observed. Exploratory analysis revealed that participants classified as cortisol responders, exhibited stronger behavioral PIT effects following acute stress compared to non-responders (Pavlovian stimulus value × stress condition × cortisol responder: estimate = −122.01, *t* = −2.29, *p* = .022).

**Figure 4.**
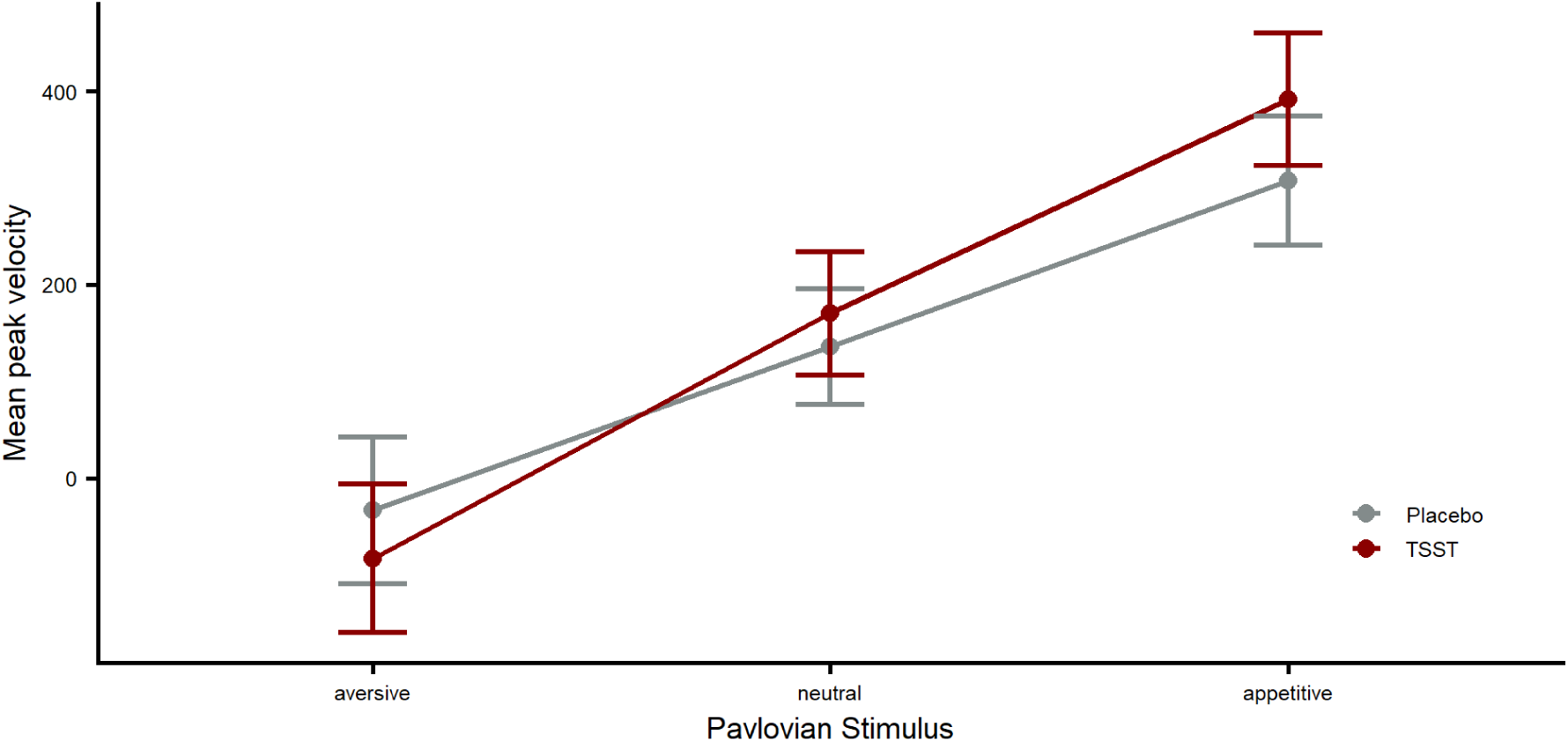
PIT Motivational. Mean peak velocity of joystick movement during monetary trials across both Controls (CON) and participants with Alcohol Use Disorder (AUD) plotted separately for stress (TSST, red) and no-stress (Placebo, gray) intervention. Although the significant interaction between stress and Pavlovian stimulus cannot be directly indicated by a significance marker (*), the effect is reflected in the steeper regression line under TSST compared with Placebo.

**Figure 5.**
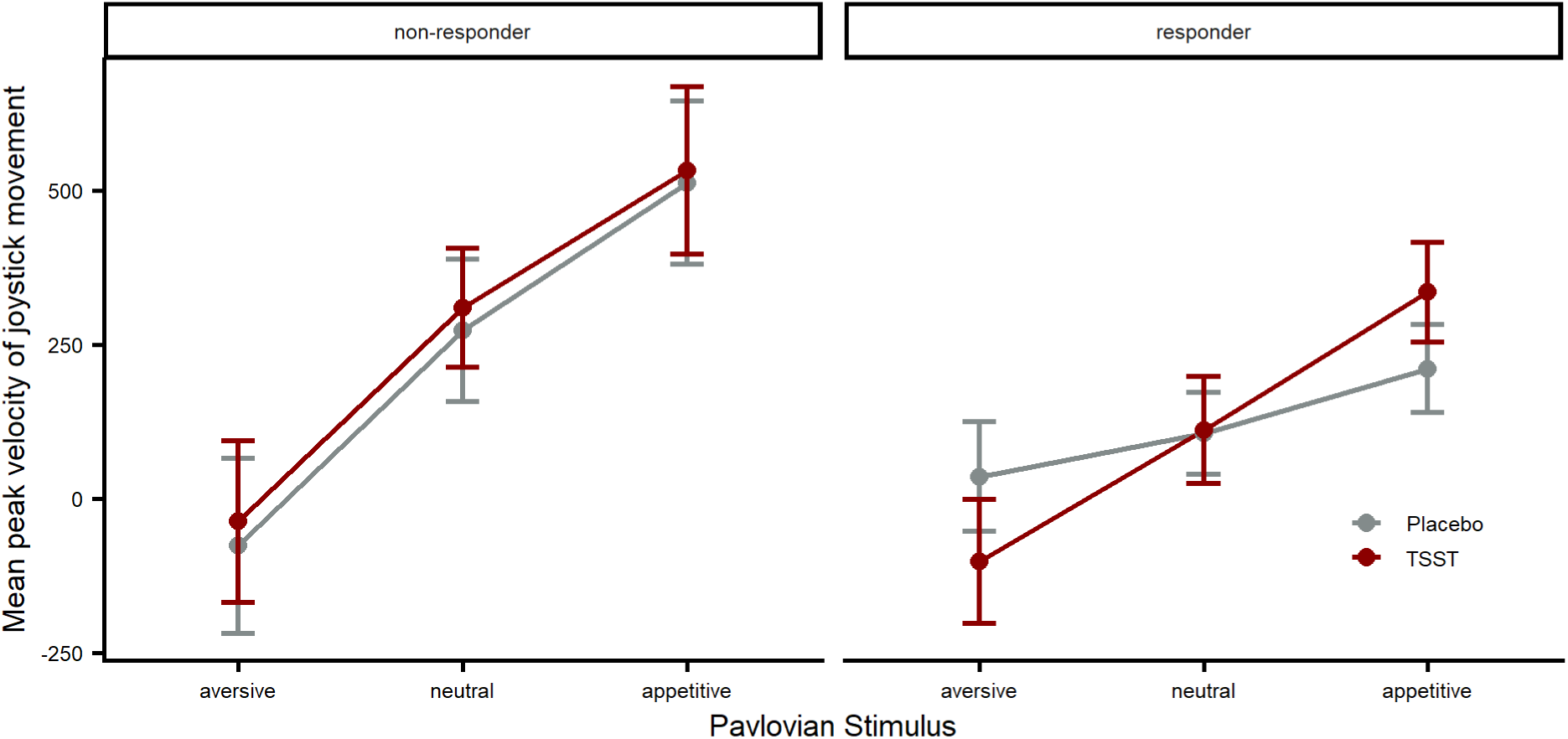
PIT Motivational Cortisol-Responder vs. Non-Responder. Mean peak velocity of joystick movement during monetary trials in cortisol non-responder (left) and cortisol-responder (right) plotted separately for stress (TSST, red) and no-stress (Placebo, gray) intervention. Regarding significant interactions, see description Figure 4.

### Stress-Dependent Modulation of PIT-Related BOLD Responses 193

Across both sessions, PIT-related activity in the right amygdala reached trend-level significance across the full sample (peak [24, −8, −12], Z = 3.13, p_FWE_ = .057; Table X). When restricting this analysis to the placebo session, right amygdala PIT-related activity reached significance (Z = 3.18, p_FWE_ = .049). During the stress session, and for the stress>placebo contrast, no significant or trend-level PIT-related activity was observed in either amygdala across the full sample (all p_FWE_ > .18; Table X).

Group comparisons (AUD>CON) revealed no significant differences during the placebo session or across both sessions combined in either hemisphere (all p_FWE_ > .68). Critically, for the stress-over-placebo contrast, AUD showed significantly greater PIT-related left amygdala activity relative to CON (peak [−24, −10, −18], Z = 3.50, p_FWE_ = .019). Given that PIT-related left amygdala activity decreased under stress relative to placebo in both groups (see Figure 6), this group difference reflects an attenuated stress-induced downregulation in AUD rather than elevated reactivity in an absolute sense. No significant nucleus accumbens activation was detected in any contrast after FWE correction, likewise the analysis of the brain-behavior relationship showed no correlation (Supplement).

**Figure 6.**
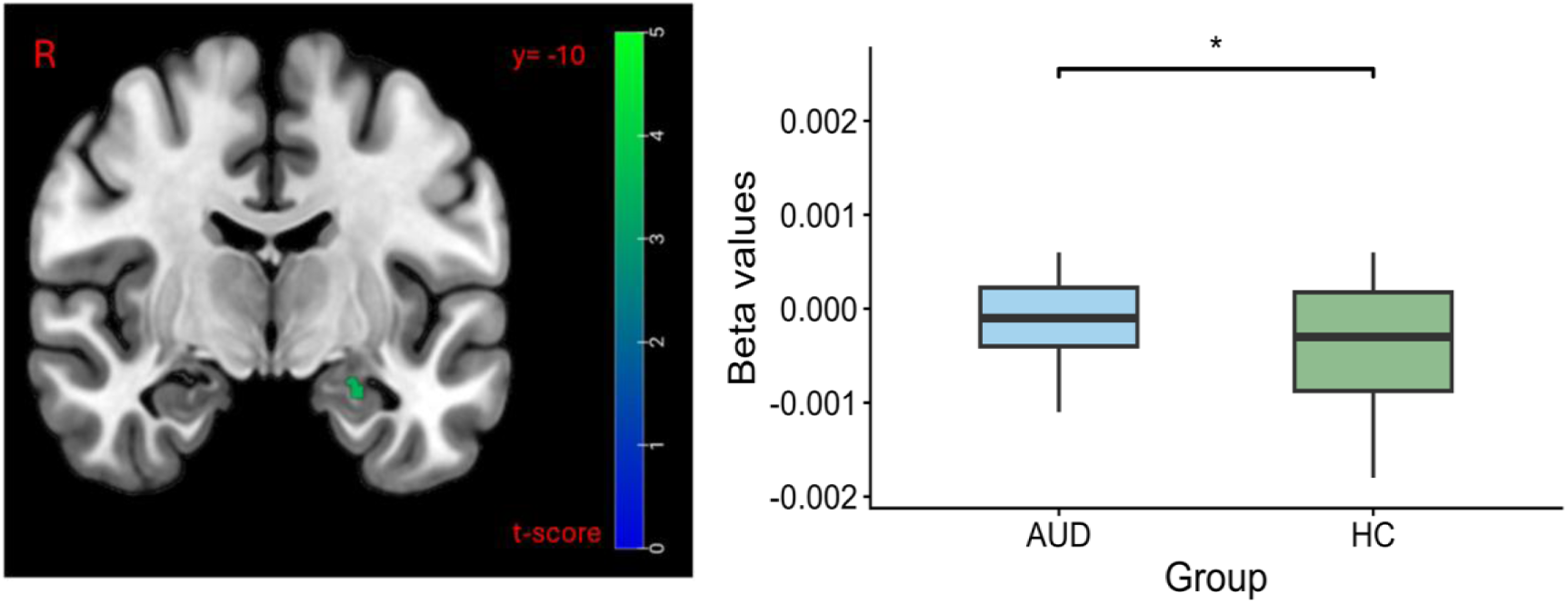
Stress-dependent Modulation of PIT-related Amygdala Activity. Group-level statistical map (left) showing a significant interaction between group (AUD > CON) and intervention (Stress > Placebo) on the PIT parametric modulator signal in the left amygdala (p < .05, FWE-corrected within the a priori compound ROI). Extracted mean beta values for individual participants are displayed on the right. Compared to controls, individuals with AUD exhibited a significantly stronger stress-induced increase in amygdala activity related to PIT (Welch’s t(54.86) = 2.15, p = .036, Cohen’s d = 0.55). Statistical tests on extracted values were conducted in R.

## DISCUSSION

The present study examined whether acute psychosocial stress alters the motivational influence of conditioned cues on instrumental behavior in individuals with AUD and CON. Three findings are particularly relevant. First, motivational PIT was robustly expressed across participants, and acute stress enhanced cue-driven instrumental response vigor, with no evidence for a differential behavioral effect in AUD. Second, exploratory analyses indicated that this stress-related potentiation was more pronounced in cortisol responders. Third, despite the absence of a behavioral group difference, stress differentially modulated PIT-related amygdala activity: PIT-related left amygdala activity decreased under stress in both groups, but this reduction was less pronounced in individuals with AUD. Together, these findings indicate that acute psychosocial stress enhances the motivational influence of conditioned cues at the behavioral level, while AUD may be associated with altered neural regulation of cue-driven processing under stress.

### Acute psychosocial stress enhances motivational PIT

The primary behavioral finding supports the hypothesis that acute stress increases PIT. Importantly, this effect was observed with non-drug-related cues. Acute stress may therefore amplify a more general process through which learned environmental cues influence ongoing behavior rather than exclusively enhancing responses to drug-associated stimuli. This is consistent with prior experimental evidence suggesting that activation of stress-related neurobiological systems increases the motivational influence of reward-predictive cues [19]. It also accords with human studies showing that acute stress can selectively enhance cue-triggered “wanting” without altering hedonic “liking” and increase the influence of Pavlovian cues on instrumental behavior [20, 21]. In the framework of incentive-sensitization theory [5, 46], this pattern is compatible with an increased motivational impact of conditioned cues under stress, although the present PIT measure does not directly quantify incentive salience or subjective “wanting”. This interpretation is in line with the conceptual framework outlined in the Introduction, in which stress and Pavlovian cues are not necessarily independent influences on behavior. Acute stress has been shown to shift behavioral control away from flexible, goal-directed processes and toward more automatic or stimulus-dependent responding [17, 18]. Pavlovian cues may similarly bias action by prioritizing previously rewarded options and thereby reducing the complexity of instrumental choice [47]. In CON, stronger PIT has previously been associated with a greater tendency toward habitual rather than complex goal-directed decision-making [48]. Under stress, the relative influence of conditioned cues on instrumental behavior may therefore increase. However, PIT should not itself be equated with habitual behavior: the present data demonstrate increased Pavlovian influence on instrumental response vigor under stress but do not establish that instrumental responding became habitual or insensitive to its consequences.

Our findings differ from Steins-Loeber et al., who found no effect of acute stress on PIT in smokers [25]. Several differences may contribute to these divergent findings. Their study examined tobacco use disorder, for which heightened PIT has not been consistently demonstrated, whereas the present study investigated AUD, in which PIT has repeatedly been associated with disease-related and prospective clinical measures [9]. In addition, Steins-Loeber et al. employed the socially evaluated cold pressor test (SECPT), whereas the present study used the TSST, which has been suggested to elicit a stronger HPA axis response [49].

PIT paradigms also vary considerably in their structure and operationalization, which further limits direct comparison [25, 50]. In exploratory analyses, cortisol responders showed stronger stress-dependent changes in PIT than non-responders. This is in line with recent reports that HPA-axis responsivity determined whether stress enhanced PIT and translated into problematic shopping behavior [51]. However, the analysis was exploratory and should therefore only be regarded as hypothesis-generating. Importantly, no group differences in motivational PIT emerged across sessions, and acute stress did not selectively amplify PIT in individuals with AUD. Thus, we did not replicate previous reports of stronger PIT in AUD [10]. Several methodological differences may account for this. In particular, previous studies from our group found that group differences were largely driven by trials requiring inhibition (“no-go”) of an instrumental response [52, 53], a condition that was not included in the present paradigm, which focused on PIT indexed by response vigor. The two-session design, with weaker PIT effects on the second day, and insufficient statistical power may have further reduced sensitivity to detect group differences. Nevertheless, the absence of an AUD-specific behavioral effect does not preclude clinical relevance. Previous studies have associated PIT with AUD, risky alcohol consumption, alcohol approach tendencies, and prospective relapse [10–14]. A general stress-induced increase in the motivational influence of conditioned cues may interact with the established vulnerability to cue-driven alcohol seeking in AUD, particularly when stress and alcohol-associated cues coincide.

### Stress-dependent modulation of PIT-related amygdala activity in AUD

At the neural level, stress altered PIT-related amygdala responses differently in individuals with AUD and CON. In the placebo condition, PIT was associated with right amygdala activation, broadly consistent with the established involvement of the amygdala in Pavlovian influences on instrumental behavior [11, 42]. No group differences emerged during the placebo day. In the left amygdala, PIT-related activity decreased under stress relative to placebo in both groups, but this reduction was less pronounced in individuals with AUD than in CON. This neural finding requires a more cautious interpretation than the behavioral effect. The present analysis demonstrates a group difference in the stress-dependent modulation of PIT-related amygdala activity; it does not directly establish impaired amygdala regulation, increased bottom-up signaling, or deficient prefrontal control in AUD.

Nevertheless, the finding is compatible with broader evidence that AUD is associated with altered adaptation of corticolimbic systems to stress [54]. Neurobiological models of addiction emphasize progressive changes in interacting reward and stress systems, including the amygdala and extended amygdala [54–56]. Recent work has also suggested reduced flexibility of orbitofrontal-amygdala connectivity during acute stress in AUD [57]. Such findings provide a possible framework for understanding why the neural processing underlying Pavlovian influences on instrumental behavior might differ under stress in AUD even when behavioral effects remain comparable between groups. Because functional connectivity was not assessed in the present analysis, however, altered prefrontal-amygdala regulation remains one possible hypothesis for future investigation rather than an explanation established by the current findings.

Notably, PIT-related activity in the placebo condition was observed in the right amygdala, whereas the stress-related group difference emerged in the left amygdala. Previous work suggests that the right amygdala mediates rapid detection and habituates quickly, while the left amygdala supports sustained evaluation of motivational information and is more often reported as active in neuroimaging research [58]. Higher exposure to daily stressors has been linked to a functional decoupling between the left amygdala and prefrontal regions like the right dorsolateral prefrontal cortex and lateral orbitofrontal cortex [59]. Concerning the role of the left amygdala in PIT, our previous research found that left amygdala activity during Pavlovian conditioning was positively associated with the strength of the subsequent behavioral PIT effect specifically in AUD [60]. However, hemispheric lateralization was not an a priori hypothesis of the present study, and replication is necessary before attributing distinct functional roles to these findings.

Crucially, neural differences were not accompanied by corresponding behavioral differences between groups and should therefore not be interpreted as evidence of stronger motivational PIT in AUD under stress. Rather, comparable behavioral effects may be accompanied by differences in underlying neural processing. Whether the observed stress-dependent amygdala difference becomes behaviorally or clinically relevant with greater AUD severity or repeated stress exposure remains an open question.

### Limitations

Several limitations should be considered. First, the behavioral sample was substantially larger than the neuroimaging sample, and some participants completed PIT outside the scanner because of limited scanning availability. Behavioral and neural analyses therefore differed in statistical power, which may have contributed to differences between effects observed at the two levels. Second, Bélanger et al. reported limited reliability for neural measures derived from a related PIT paradigm [26]. Although the present task and modelling approach differ in relevant respects [29], the current neural findings should therefore be considered in need of independent replication. Third, despite previous evidence linking the NAcc to motivational PIT and relapse in AUD [12, 13], we did not detect significant PIT-related NAcc activation.

NAcc analyses were constrained by comparatively lower fMRI signal quality in this region than in the amygdala (Supplementary Material), reducing sensitivity to detect PIT-related effects. Accordingly, the absence of significant NAcc activation cannot be taken as evidence against its involvement in motivational PIT. Fourth, we found no significant associations between extracted neural estimates and individual behavioral PIT slopes, however this should be interpreted cautiously, particularly given the limited reliability of brain-behavior correlations in moderate neuroimaging samples [61]. Finally, interference-PIT analyses are reported in the Supplementary Material. Although these analyses showed broadly similar stress-related patterns, interference PIT represents a distinct operationalization and was assessed only as a complementary measure

### Conclusions

Acute psychosocial stress enhanced the motivational influence of Pavlovian cues on instrumental behavior across individuals with AUD and CON. These findings identify PIT as one potential process through which stress may increase alcohol-seeking behavior. Behaviorally, the PIT effect was not specific to AUD. At the neural level, however, individuals with AUD and CON differed in the stress-dependent modulation of PIT-related amygdala activity. Given the absence of corresponding behavioral group differences and the lack of direct measures of regulatory connectivity, the functional significance of this neural difference remains to be established. Future longitudinal studies should determine whether stress-related changes in motivational PIT and its neural correlates predict clinically meaningful outcomes, including loss of control and relapse in AUD.

## Data Availability

All data produced in the present study are available upon reasonable request to the authors

