## Supplementary Material for "Acute Social Stress Potentiates Cue-Driven Behavior in Alcohol Use Disorder: Behavioral and Neural Evidence"

**Overview:**

**S1 Flowchart**

**S2 Participant inclusion and exclusion criteria**

**S3 Demographic analysis**

**S4 PIT Paradigm**

**S5 fMRI data acquisition and preprocessing**

**S6 fMRI data and quality control procedures**

**S7 Design matrix (Motivational PIT)**

**S8 PIT Interference**

**S9 Salivary cortisol analysis**

**S10 Subjective Analysis**

**S11 Behavioral Analysis and Results**

**S12 Motivational PIT: Random-Effect Structure**

**S13 Motivational PIT: Detailed Results and Exploratory PIT Cortisol Analysis**

### S1 Flowchart

#### Figure S1: Participant Flowchart

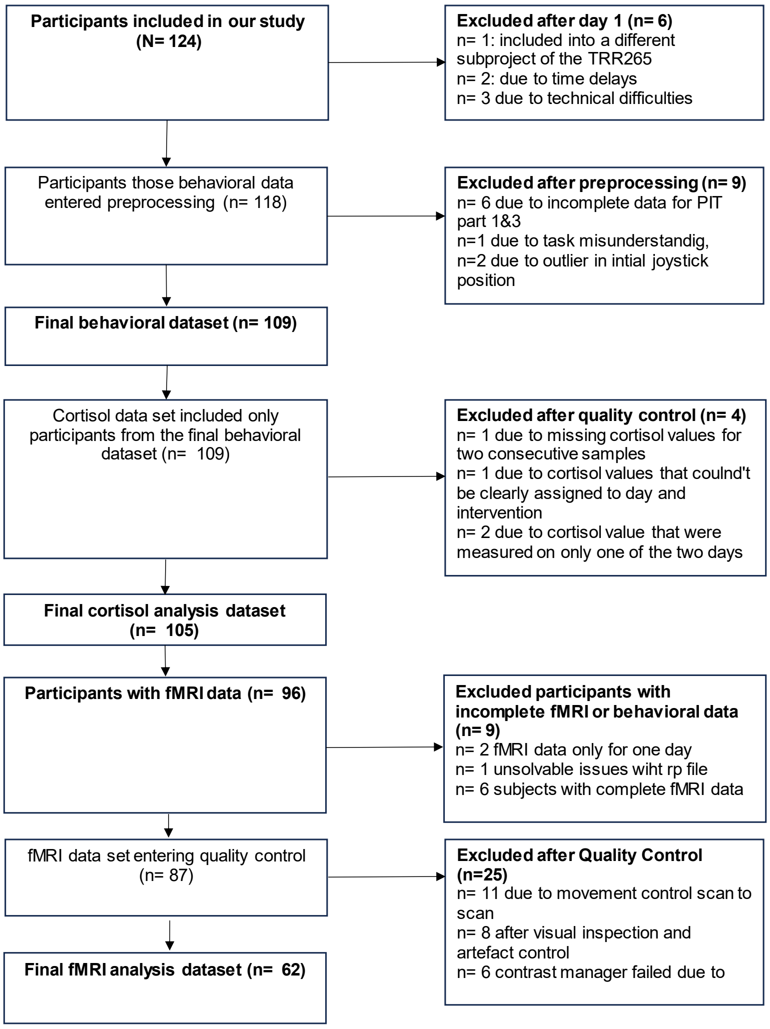

### S2 Participant inclusion and exclusion criteria

**Inclusion criteria.** Participants were eligible if they (1) were men or women aged 16–32, 33–49, or 50–65 years; (2) met DSM-5 criteria for mild to moderate alcohol use disorder (AUD; 2 to a maximum of 5 fulfilled criteria) and did not clinically require detoxification, as confirmed by an independent board-certified psychiatrist - AUD patients with comorbid mild to moderate cannabis use disorder and/or tobacco use disorder were eligible; (3) were able to provide fully informed consent and to complete self-rating scales; (4) were willing to use an Android smartphone; and (5) had sufficient understanding of the German language.

**Exclusion criteria.** Participants were excluded if they had (1) a lifetime history of DSM-5 bipolar disorder, schizophrenia or a schizophrenia-spectrum disorder, or substance dependence other than alcohol, nicotine, or cannabis dependence (severe alcohol or cannabis use disorder was also exclusionary); (2) a current threshold DSM-5 diagnosis of major depressive disorder, or presence of suicidal intention; (3) a history of severe head trauma or another severe central nervous system disorder (e.g., dementia, Parkinson's disease, multiple sclerosis); (4) pregnancy or nursing an infant; or (5) current use of medication(s) or drug(s) known to interact with the central nervous system, unless at least four half-lives had elapsed since the last intake.

### S3 Demographic Analysis

A more extensive sample characteristics can be found below in Table S1.

#### Table S1. Sample Characteristics

| Characteristic | n | HC | n | AUD | p |
|---|---|---|---|---|---|
| **SOCIODEMOGRAPHIC VARIABLES** | | | | | |
| Female, n (%) | 46 | 29 (63.0%) | 63 | 29 (46.0%) | 0.118[a] |
| Age, years | 46 | 35.09 (11.48) | 63 | 39.46 (12.38) | 0.060[c] |
| Education, years | 44 | 12.55 (0.90) | 59 | 12.68 (0.92) | 0.466[c] |
| Smokers, n (%) | 46 | 5 (10.9%) | 62 | 26 (41.9%) | <.001[a] |
| **Monthly net income bracket** | 46 | | 62 | | 0.851[b] |
| <500 | | 5 (10.9%) | | 5 (8.1%) | |
| 500-1000 | | 5 (10.9%) | | 8 (12.9%) | |
| 1000-1500 | | 6 (13.0%) | | 6 (9.7%) | |
| 1500-2000 | | 8 (17.4%) | | 10 (16.1%) | |
| 2000-2500 | | 8 (17.4%) | | 7 (11.3%) | |
| 2500-3500 | | 10 (21.7%) | | 12 (19.4%) | |
| 3500-4000 | | 3 (6.5%) | | 6 (9.7%) | |
| 4000-4500 | | 1 (2.2%) | | 4 (6.5%) | |
| 4500-5000 | | 0 (0.0%) | | 1 (1.6%) | |
| >5000 | | 0 (0.0%) | | 3 (4.8%) | |
| **AUD SEVERITY** | | | | | |
| AUD criteria, last 12mo | 26 | 0.12 (0.33) | 63 | 4.37 (1.95) | <.001[c] |
| AUD criteria, lifetime | 26 | 1.04 (1.87) | 63 | 4.51 (2.32) | <.001[c] |
| AUDIT sum | 46 | 2.98 (2.59) | 62 | 15.45 (6.46) | <.001[c] |
| Craving (CAS-A total score) | 46 | 1.20 (2.67) | 62 | 18.42 (11.62) | <.001[c] |
| **ALCOHOL CONSUMPTION PATTERN (PAST 3 MONTHS)** | | | | | |
| Drinking days | 45 | 7.68 (8.71) | 63 | 52.75 (24.03) | <.001[c] |
| Usual amount per drinking day | 45 | 2.27 (1.90) | 63 | 5.85 (2.69) | <.001[c] |
| Est. weekly volume (derived) | 45 | 0.79 (1.17) | 63 | 21.95 (15.34) | <.001[c] |
| **PSYCHIATRIC COMORBIDITY** | | | | | |

| Characteristic | n | HC | n | AUD | p |
|---|---|---|---|---|---|
| Depressivity (ADS-K total score) | 46 | 6.85 (4.71) | 62 | 10.26 (5.74) | <.001[c] |
| **Panic disorder status** | 46 | | 63 | | 1.000[b] |
| never | | 46 (100.0%) | | 63 (100.0%) | |
| past | | 0 (0.0%) | | 0 (0.0%) | |
| current | | 0 (0.0%) | | 0 (0.0%) | |
| **Social anxiety disorder status** | 46 | | 63 | | 1.000[b] |
| never | | 46 (100.0%) | | 62 (98.4%) | |
| past | | 0 (0.0%) | | 0 (0.0%) | |
| current | | 0 (0.0%) | | 1 (1.6%) | |
| **GAD status** | 36 | | 0 | | 1.000[b] |
| never | | 36 (100.0%) | | 0 (-) | |
| past | | 0 (0.0%) | | 0 (-) | |
| current | | 0 (0.0%) | | 0 (-) | |
| Trait anxiety (STAI-T total score) | 0 | - | 62 | 39.35 (9.62) | |
| **PERSONALITY** | | | | | |
| Impulsivity (BIS-15 total score) | 46 | 29.61 (6.51) | 62 | 32.81 (6.83) | 0.015[c] |
| **NEUROCOGNITIVE FUNCTIONING** | | | | | |
| Verbal intelligence (WST) | 46 | 27.80 (6.81) | 63 | 29.10 (7.36) | 0.347[c] |
| Working memory (Digit Span Backward) | 46 | 5.20 (1.31) | 63 | 4.94 (1.11) | 0.279[c] |
| Fluid intelligence (Matrices) | 44 | 6.07 (3.14) | 63 | 6.17 (2.63) | 0.854[c] |

Note. Continuous variables reported as M (SD); categorical as n (%). Education in years is an approximation from highest school degree (Abitur coded as 13 years).

[a] Chi-square test. [b] Fisher's exact test. [c] Welch's two-sample t-test. Trait anxiety (STAI-T) assessed in the AUD group only.

### S4 PIT Paradigm

#### Part 1: Instrumental Learning

Participants learned to collect or reject shells or leafs with a joystick, contingent upon whether the presented stimuli were classified as "good" or "bad." The collection (i.e. the pulling with the joystick) of a "good" shell or leaf was rewarded 80% of the time with a gain of 20 cents, while the remaining 20% resulted in a loss of the same amount. Conversely, the rejection (i.e. the pushing away with the joystick) of a “bad" shell or leaf led to a reward in 80% of cases and a loss of 20 cents in the remaining 20%. Immediate feedback, in the form of visual cues indicating monetary gain or loss followed each response. On one of the two experimental days instrumental stimuli were shells, on the alternate day participants were presented leafs. The classification of stimuli and their presentation order were randomized across participants to ensure experimental control. The instrumental training concluded either upon achieving an 80% correct choice criterion over 16 trials or after completing a maximum of 120 trials. Participants were required to complete a minimum of 60 trials to ensure comparable success in instrumental learning across participants.

#### Part 2: Pavlovian conditioning

During Pavlovian training, five abstract fractals paired with auditory tones (CS) were associated with either monetary outcomes (appetitive: +10€, neutral: 0€, aversive: −10€) or drug-related images (alcohol, smoking) as unconditioned stimuli (US). Each trial began with the presentation of a compound CS (fractal picture and pure tone) for 3 s, followed by a 3 s delay and then the US (monetary outcome or drug-related image) appeared for 3 s. The task design included one appetitive condition (CS paired with monetary gain), one aversive condition (CS paired with monetary loss) and a neutral condition (no monetary gain or loss). In addition, two drug-related conditions were incorporated, representing a novel extension of our group’s earlier paradigms, which had employed a broader range of monetary conditions and smoking- and alcohol pictures  (Garbusow et al.; Ebrahimi et al.) but did not include drug-related CS. Each of the five CS was presented 16 times, yielding a total of 80 trials. Participants were instructed to pay attention to and memorize the CS- US associations.

**Part 3: Pavlovian to instrumental transfer**

In this phase of the experiment, participants were instructed to repeat the task from part 1, collecting “good” shells or leafs and rejecting “bad” ones, while the Pavlovian CS were displayed in the background. Unlike during the instrumental learning part, no immediate feedback was provided, meaning the task was performed under nominal extinction. Participants were informed that monetary gains accumulated during the task would be paid out at the end of the session and that the Pavlovian CSs were irrelevant for task performance. Across 180 trials, each one of the five Pavlovian CS appeared 36 times whereof 6 time with each one of the 6 different instrumental stimuli.

**Part 4: Query trials**

Participants completed a forced-choice task designed to assess the implicit contingency awareness. In each trial, a pair of two CSs was presented: first individually for 1 s, followed by a display of both CSs side by side for up to 2 s. Participants were instructed to select the “more appealing” CS. Each possible CS pair was presented three times in randomized order.

### S5 fMRI data acquisition and preprocessing

Structural and functional MRI data acquisition was conducted on 3-Tesla Siemens Trio scanners (Siemens AG, Erlangen, Germany) equipped with a 64-channel head coil at the Berlin Center for Advanced Neuroimaging Core Facility (RRID:SCR_028020, https://www.berlin-can.de/). Functional images were collected using a T2*-weighted multiband echo-planar imaging (EPI) sequence (TR = 869 ms, TE = 38 ms, flip angle = 58°, matrix = 88 × 88, FOV = 210 mm, 60 slices, voxel size = 2.4 × 2.4 × 2.4 mm, multiband factor = 6). For anatomical reference, a high-resolution T1-weighted structural image was obtained (TR = 2000 ms, TE = 2.01 ms, flip angle = 8°, matrix = 256 × 256, FOV = 256 mm, 208 slices, 1 mm isotropic resolution). Additionally, a dual-echo gradient field map was acquired for correcting geometric distortions in the EPI images.

Data preprocessing included standard steps: slice timing correction, realignment to the mean functional image and distortion correction based on fieldmaps (unwarping). The T1-weighted image was segmented and the mean functional image was co-registered to the resulting tissue maps. Data were then normalized to MNI space using a 2 mm isotropic voxel resolution and spatially smoothed with a Gaussian kernel (6 mm FWHM). A high-pass temporal filter (cutoff: 128 s) was applied prior to ,first-level analysis.

### S 6 fMRI data and quality control procedures

#### Visual inspections and incidental findings

For each participant, we manually inspected the MRI data to identify and exclude artefacts, such as signal inhomogeneity and loss, ghosting or spiking. Our procedure was guided by the recommendations of the Centre for Brain Science, Massachusetts General Hospital and Harvard University (MRI Quality Control guidelines: https://cbs.fas.harvard.edu/facilities/neuroimaging/investigators/mr-data-quality-control).

Feldfunktion geändert

#### Quality Control of Regions of Interest (ROI)

To verify adequate data quality in all ROIs (separately for bilateral Amygdala, left Nucleus accumbens and right Nucleus accumbens), we ran a function in Matlab ("ROIana") previously established by our work group to extract voxelwise activity within probabilistic ROI masks (Chen et al., 2023). For each participant, we quantified the number of voxels included after applying different probability thresholds (0.2–0.8) to the masks and compared this to the total voxel count of the ROI. Participants with <90% coverage in a given ROI were considered as failing quality control. For the final analyses, a probability threshold of 0.2 was selected. For the bilateral amygdala, voxel coverage remained stable across thresholds up to 0.4, with no participants excluded until a very conservative threshold of 0.8, where 20 participants would have been removed. In contrast, left and right Nucleus accumbens showed strongly reduced coverage at stricter thresholds: already at 0.3, 7 participants fell below the 90% criterion, increasing to 18–20 at 0.4 and >60 at 0.8. Thus, while the amygdala ROIs were robust to thresholding, both the right and the left Nucleus Accumbens ROI were highly sensitive, leading to reduced effective sample sizes and potentially contributing to the absence of significant Nucleus accumbens findings.

**Head Motion Quality Control**

We applied a scan-to-scan movement control procedure. For each participant, the original realignment parameter (rp) files were extracted and framewise displacements were calculated across the six motion dimensions (x, y, z translations; pitch, roll, yaw rotations). We set a threshold of 4 mm for translations and 3° for rotations as exclusion criteria. Two participants exceeded the 3° threshold slightly (3.0xx°) but were retained, as their data did not show further artefacts.

**Second-Level Sanity Checks**

As an additional quality control measure, we performed second-level sanity checks to confirm expected activation patterns. Specifically, we inspected the contrast capturing visual responses and observed robust activation in the occipital cortex. Likewise, the contrast reflecting motor responses revealed expected activation patterns in the motor cortex.

### S7 Design matrix (Motivational PIT)

For motivational PIT, both experimental days were modelled within one design matrix as shown below in Figure S2.

**Figure S2: Design Matrix Motivational PIT**

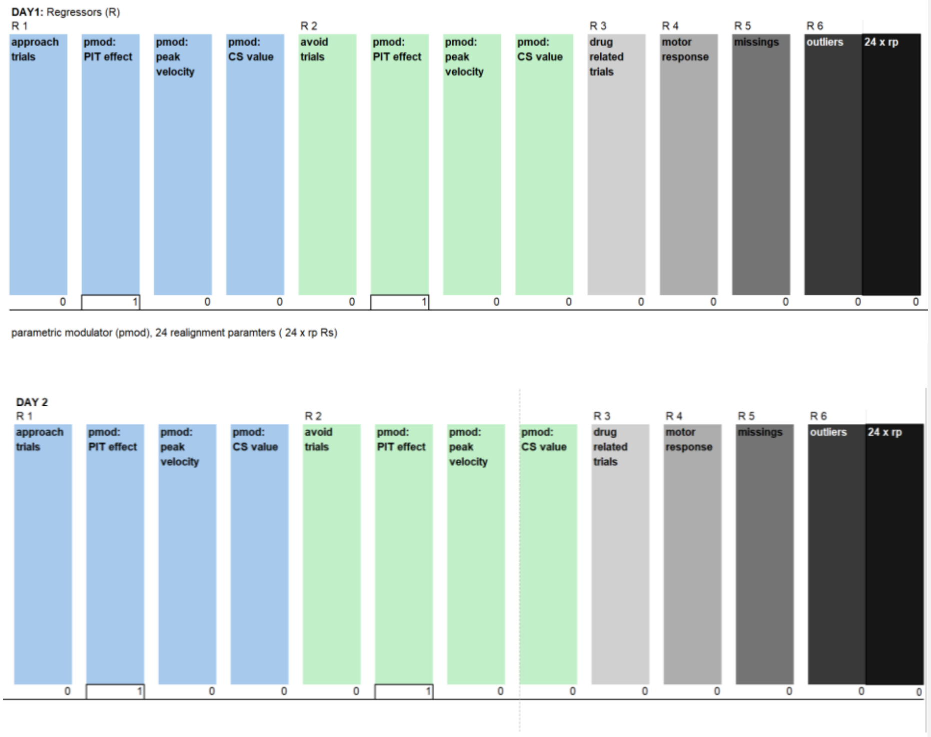

### S8 Interference PIT

To broaden our characterization of stress effects on PIT, we additionally examined PIT interference, an established approach capturing a distinct facet of PIT, namely susceptibility to interference during Pavlovian-instrumental conflict, that complements the motivational PIT analyses reported in the main text. As this analysis was not part of our primary confirmatory hypotheses, it was conducted on an exploratory basis. Therefore all reported p-values should be interpreted as descriptive rather than confirmatory.

**Interference PIT - Behavioral Analysis**

Following the approach described by Belanger et al. 2025, an individual interference score was computed as the difference in error rates (ER) between incongruent and congruent trials (ER incongruent − ER congruent) for each participant and experimental session. A t-test was conducted to confirm the presence of the basic PIT interference effect, testing whether error rates were higher on incongruent relative to congruent trials, consistent with our hypothesis that incongruent trials elicit more errors. Interference scores were subsequently averaged separately for the TSST and placebo sessions for each participant, and a paired-samples t-test compared interference scores between sessions, testing whether acute stress increased PIT interference relative to placebo. For the final group comparison, a differential score (ER stress − ER placebo) was calculated and compared between individuals with AUD and HC, testing whether the stress-induced increase in PIT interference was more pronounced in the AUD group. To examine the relationship between motivational and interference PIT, individual motivational PIT scores were obtained from the final linear mixed-effects model by combining the fixed-effect estimate for Pavlovian cue value with each participant's best linear unbiased predictor (BLUP) for the Pavlovian cue slope. These subject-specific motivational PIT scores were correlated with mean interference PIT scores across sessions using Spearman's rank correlation, as both variables deviated from normality according to Shapiro-

Wilk tests.

**Interference PIT - Behavioral Results**

Participants showed higher error rates during incongruent than congruent trials, consistent with a PIT interference effect (mean interference score = 0.102, t(219) = 6.12, p < .001). Interference scores were higher following acute stress than following placebo, consistent with our hypothesis that stress enhances PIT interference (mean difference = 0.037, t(108) = 1.83, p = .035). The stress-induced increase in interference did not differ significantly between groups (t(104.79) = −0.11, p = .545), providing no support for a more pronounced stress effect in individuals with AUD.

**Figure S3: Behavioral Results of Interference PIT**

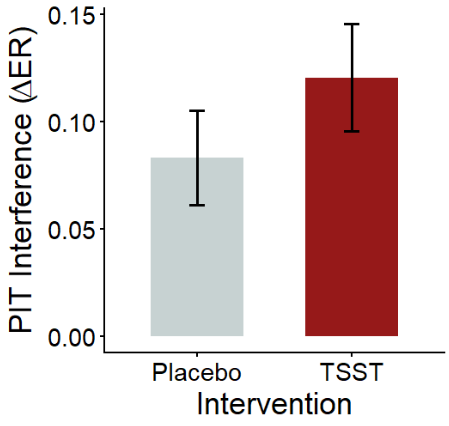


Our two indexes to measure the Pavlovian to Instrumental transfer, the motivational PIT and interference PIT were moderately positively correlated (Spearman's ρ = .56, *p* < .001), indicating that individuals showing stronger motivational PIT effects also tended to exhibit larger interference effects.

**Interference PIT - Imaging Analysis Method**

Analogously to motivational PIT, individual first-level general linear models (GLMs) were implemented in SPM12 (Wellcome Centre for Human Neuroimaging, London, UK) using an event-related design and both experimental sessions (TSST and Placebo) were modelled within a single design matrix. The first-level model comprised 11 regressors modelled as stick functions: as regressors of main interest, the three monetary Pavlovian conditioned stimuli (−10€, 0€, +10€) × the two instrumental conditions (approach and avoid), as regressors of no interest, two drug-related Pavlovian CS (alcohol, smoking) × the two instrumental conditions and one regressor capturing motor-related variance, with individual joystick movement initiation time as the event onset. All first-level models included 24 head motion parameters as nuisance regressors (six realignment parameters, their first derivatives, and their squared terms). A high-pass filter of 128 s was applied, and serial autocorrelations were modelled using an AR(1) process. At the first level, the incongruent-versus-baseline contrast was computed by collapsing the two incongruent trial types (types (CS− paired with approach and CS+ paired with avoid) and subtracting the neutral baseline condition (CS neutral paired with either instrumental response), an approach recently shown to exhibit superior test-retest reliability compared to the incongruent versus congruent contrast that was previously used (Belanger et al., 2025). To account for the counterbalanced within-subject crossover design, subject-specific weighted contrast vectors were applied to the two-session design matrix, yielding three contrasts of interest per contrast type: (1) an overall PIT contrast collapsing across both sessions, (2) a stress-over-placebo contrast (TSST > placebo), and (3) a stress-session-only contrast. Individual first-level contrast images were entered into second-level random-effects analyses. Group-level one-sample t-tests were conducted to assess whether PIT-related activation exceeded baseline across all participants. Two-sample t-tests compared activation between individuals with alcohol use disorder (AUD) and healthy controls (HC). Individual behavioral PIT interference scores were included as covariates of interest at the second level to characterise the brain-behavior relationship underlying the neural incongruence effect. Region-of-interest (ROI) analyses were performed for four a priori defined regions known to be implicated in PIT: the ventral striatum (VS), amygdala, dorsomedial prefrontal cortex (dmPFC), and lateral prefrontal cortex (lPFC), using anatomical masks defined as in previous work (Chen et al., 2021). Mean parameter estimates were extracted from each ROI using the eigenvariate approach in SPM12 without data adjustment, separately for the incongruent-versus-congruent and incongruent-versus-baseline contrasts. Extracted beta values were then subjected to one-sample t-tests to assess overall PIT-related activation, two-sample t-tests for group comparisons, and Pearson correlation tests to assess the association between ROI-averaged neural PIT responses and individual behavioral interference scores. All ROI-level tests were subjected to Bonferroni correction across the four ROIs within each contrast family, yielding a corrected threshold of p corrected = 0.0125 ($\alpha = 0.05 / 4$). For exploratory purposes, whole-brain analyses were conducted for the incongruent-versus-congruent contrast with the behavioral interference score as a covariate, applying an uncorrected voxel-level threshold of $p < .001$ with a minimum cluster extent of $k \geq 30$ contiguous voxels.

**Interference PIT - Imaging Analysis Results**

<u>Incongruent-versus-baseline contrast</u>

*PIT overall.* One-sample tests revealed a significant incongruent-versus-baseline effect in the dorsomedial prefrontal cortex (M = 18.40, t(121) = 2.59, p = .011, p corrected= .043); no other ROI reached significance. No group differences were observed in any ROI for this contrast (all p corrected> .52).

*PIT stress-over-placebo.* No ROI showed a significant one-sample or group effect after correction. The dorsomedial prefrontal cortex showed a trend-level reduction in the one-sample test prior to correction (M = −15.49, t(121) = −1.87, p = .064, p corrected= .257).

**Table S2. Incongruent-versus-Baseline Contrast: ROI Parameter Estimates**

| Contrast | ROI | One-sample M | t(df) | p | p_corr | M AUD / M HC | Group t(df) | p | p_corr |
|---|---|---|---|---|---|---|---|---|---|
| Overall | VS | 5.52 | 0.71(121) | .481 | 1.000 | 4.40 / 6.94 | −0.16(119.99) | .869 | 1.000 |
| Overall | AMY | 6.24 | 0.79(121) | .432 | 1.000 | −4.42 / 19.66 | −1.52(112.40) | .132 | .529 |
| Overall | dmPFC | 18.40 | 2.59(121) | .011 | **.043** | 18.17 / 18.70 | −0.04(119.97) | .970 | 1.000 |
| Overall | lPFC | 2.41 | 0.32(121) | .750 | 1.000 | 2.54 / 2.24 | 0.02(115.86) | .984 | 1.000 |
| Stress > Placebo | VS | −1.51 | −0.17(121) | .866 | 1.000 | −6.69 / 5.01 | −0.64(111.02) | .521 | 1.000 |
| Stress > Placebo | AMY | −4.95 | −0.50(121) | .617 | 1.000 | −9.31 / 0.55 | −0.51(119.66) | .614 | 1.000 |
| Stress > Placebo | dmPFC | −15.49 | −1.87(121) | .064 | .257 | -20.47 / -9.21 | -0.70(118.11 | .485 | 1.000 |
| Stress > Placebo | lPFC | −4.57 | −0.58(121) | .560 | 1.000 | −14.24 / 7.60 | −1.52(89.89) | .132 | .529 |

*Bold values indicate p corrected< .05 after Bonferroni correction (α = .05/4 within each contrast family). VS = ventral striatum; AMY = amygdala; dmPFC = dorsomedial prefrontal cortex; lPFC = lateral prefrontal cortex.*

### Results: PIT Interference - Whole-Brain Analysis

*PIT overall*. With the behavioral interference score included as a covariate, the incongruent-versus-congruent contrast revealed a trend-level cluster in the right gyrus rectus (peak [10, 40, −22], p < .001, pFWE = .051), approaching but not surviving whole-brain FWE correction. A second, non pFWE significant cluster was observed in the right superior frontal gyrus, medial segment (peak [4, 46, 38], p < .001, pFWE = .768).

*PIT placebo*. Within the right superior frontal gyrus medial segment a significant cluster was found (peak [2, 36, 30], p < .001, pFWE = .77], another one was detected in the left middle frontal gyrus (peak [-32, 48, 12], p < .001, pFWE = .945).

*PIT stress* No suprathreshold clusters were observed.

*PIT stress-over-placebo* No suprathreshold clusters were observed in either the one-sample or two-sample (group comparison) whole-brain analyses

### S9 Salivary cortisol analysis

#### Exclusion criteria, missing data and imputation

Participants were excluded from the cortisol analysis if they met any of the following criteria: (1) missing cortisol values for two consecutive samples, (2) cortisol values available for only one of the two experimental days or (3) cortisol values that could not be clearly attributed to the TSST or Placebo-TSST intervention. Four participants had to be excluded, therefore 105 of the 109 participants with complete behavioral data were included into the cortisol analysis. Imputation was performed by substituting the T6 value for a missing T7 value and the T2 value for a missing T1 value. Missing values between T2 and T6 were not imputed. For participants with such missing data, AUCg was computed individually. This imputation logic was applied to a total 6 of participants with a missing value.

**Table S3. Repeated Measures ANOVA for AUCg**

| Effect | Df num | Df den | F | p | $\eta^2$ | Significance |
|---|---|---|---|---|---|---|
| (Intercept) | 1.00 | 103.00 | 472.83 | < .001 | 0.78 | * |
| Group (AUD, HC) | 1.00 | 103.00 | 0.08 | 0.783 | <0.001 | |
| Intervention | 1.00 | 103.00 | 51.29 | < .001 | 0.11 | * |
| Group x Intervention | 1.00 | 103.00 | 0.22 | 0.642 | <0.001 | |

**Table S4. Repeated Measures ANOVA for Baseline cortisol**

| Effect | Df num | Df den | F | p | η² | Significance |
|---|---|---|---|---|---|---|
| (Intercept) | 1.00 | 103.00 | 345.28 | < .001 | 0.70 | * |
| Group (AUD, HC) | 1.00 | 103.00 | 0.59 | 0.443 | <0.001 | |
| Intervention | 1.00 | 103.00 | 0.07 | 0.793 | <0.001 | |
| Group x Intervention | 1.00 | 103.00 | 0.19 | 0.662 | <0.001 | |

### S10 Subjective Analysis

Due to missing data, two participants were excluded, yielding a final sample of N = 109.

12886 had a missing for day 1: +85, 12869 had a missing for day 2: -10

**Table S5. Repeated measures ANOVA on peak arousal**

| Effect | df um | Df den | F | p | η² | Significance |
|---|---|---|---|---|---|---|
| (Intercept) | 1.00 | 104.00 | 88.70 | < .001 | 0.32 | * |
| Group (AUD, HC) | 1.00 | 104.00 | 0.57 | 0.451 | 0.00 | |
| Intervention | 1.00 | 104.00 | 32.87 | < .001 | 0.13 | * |
| Group x Intervention | 1.00 | 104.00 | 2.25 | 0.137 | 0.01 | |

**Table S6. Repeated measures ANOVA on peak stress**

| Effect | Df num | Df den | F | p | η² | Significance |
|---|---|---|---|---|---|---|
| (Intercept) | 1.00 | 104.00 | 137.88 | < .001 | 0.38 | * |
| Group (AUD, HC) | 1.00 | 104.00 | 1.02 | 0.314 | 0.00 | |
| Intervention | 1.00 | 104.00 | 100.72 | < .001 | 0.34 | * |
| Group x Intervention | 1.00 | 104.00 | 2.36 | 0.128 | 0.01 | |

**Table S7. Repeated measures ANOVA on peak valence**

| Effect | Df num | Df den | F | p | η² | Significance |
|---|---|---|---|---|---|---|
| (Intercept) | 1.00 | 104.00 | 140.02 | < .001 | 0.38 | * |
| Group (AUD, HC) | 1.00 | 104.00 | 1.84 | 0.178 | 0.01 | |
| Intervention | 1.00 | 104.00 | 82.03 | < .001 | 0.30 | * |

| Effect | Df num | Df den | F | p | η² | Significance |
|---|---|---|---|---|---|---|
| Group x Intervention | 1.00 | 104.00 | 0.22 | 0.636 | 0.00 | |

**Subjective Stress Responses by Cortisol Responder Status**

To examine whether subjective responses to the stress manipulation differed as a function of cortisol responder status, separate repeated-measures ANOVAs were conducted for peak arousal, peak stress, and peak valence, with intervention (TSST vs. placebo) as the within-subject factor and cortisol responder status (responder vs. non-responder) as the between-subject factor. Across all three subjective measures, significant main effects of intervention indicated robust changes following the TSST. Peak arousal ($F(1, 104) = 10.62$, $p = .002$, $\eta^2_p = .09$), peak stress ($F(1, 104) = 28.89$, $p < .001$, $\eta^2_p = .22$), and peak valence ($F(1, 104) = 30.71$, $p < .001$, $\eta^2_p = .23$) differed between the TSST and placebo conditions. No significant main effects of cortisol responder status were observed for peak arousal ($F(1, 104) = 0.57$, $p = .453$), peak stress ($F(1, 104) = 3.26$, $p = .074$), or peak valence ($F(1, 104) = 0.32$, $p = .571$). Importantly, none of the responder status × intervention interactions reached significance for peak arousal ($F(1, 104) = 0.02$, $p = .880$), peak stress ($F(1, 104) = 0.50$, $p = .482$), or peak valence ($F(1, 104) = 0.03$, $p = .873$), indicating that subjective responses to the TSST did not significantly differ between cortisol responders and non-responders.

### S11 Behavioral PIT Analysis: Quality Control, Instrumental and Pavlovian Learning

**Quality Control and Exclusion Criteria**

To ensure data quality for the behavioral PIT tasks (Parts 1 and 3), we examined missed responses and outliers in initial joystick position. Across all monetary PIT trials on both days,

0.92% of responses were missings. During instrumental learning, 1.36% of trials were missed. For analysis, both missing trials and trials with outliers in initial joystick position were excluded from PIT Part 3. In contrast, for Part 1, outlier trials were retained since performance was assessed by accuracy rather than peak velocity.

To control for outliers related to initial joystick position during PIT, trials were excluded when the joystick displacement at trial onset exceeded a symmetric ±0.28-unit cutoff. This threshold was chosen based on its symmetry around the neutral position and because it approximately corresponded to a ±3 SD range in the present dataset. Importantly, peak velocity was computed independently of the initial joystick position; therefore the exclusion criterion was chosen conservatively to remove trials with clearly displaced starting positions. Participants were excluded if more than 40% of trials on either testing day exceeded this cutoff, resulting in the exclusion of two participants. For the remaining sample, outlier trials were excluded at the trial level. Across the full dataset 0.93% of trials were excluded due to initial joystick position. Missing and outlier trials were modeled as separate regressors of no interest in the imaging analysis.

**Task misunderstanding.** One participant was excluded for task misunderstanding, defined as exclusive reliance on Pavlovian cues, indicated by a mean error rate of 1 in incongruent trials and 0 in congruent trials, combined with a debriefing report that the participant believed Pavlovian cues were informative for shell quality. Although three participants met the behavioral criterion, only one additionally fulfilled the debriefing criterion and was therefore excluded.

**Instrumental Learning**

Three-way ANOVAs were conducted to assess instrumental learning, examining the effects of group (AUD, HC), day (day 1, day 2), and intervention (stress, no-stress) on mean accuracy over the last 16 trials. Participants demonstrating performance significantly above chance level during the last 16 trials, as determined by a chi-square test, were classified as learners.

The three-way ANOVA for mean accuracy over the last 16 trials revealed a significant main effect of day, $F(1, 210) = 26.35$, $p < .001$, indicating differences in instrumental learning performance between the two days of testing. In addition, a significant day × intervention interaction emerged, $F(1, 210) = 6.38$, $p = .012$, suggesting that changes in mean accuracy between days differed depending on the intervention. No significant main effects of group, $F(1, 210) = 0.61$, $p = .436$, or intervention, $F(1, 210) = 0.50$, $p = .479$, were observed. Likewise, neither the day × group interaction, $F(1, 210) = 0.01$, $p = .905$, the group × intervention interaction, $F(1, 210) = 0.10$, $p = .751$, nor the three-way day × group × intervention interaction, $F(1, 210) = 0.08$, $p = .778$, reached significance. On the first day, 82 out of 109 participants (75.23%) achieved learner status, while on the second day, 102 out of 109 participants (93.58%) performed significantly above chance level during the last 16 trials. Within the AUD group, 48 out of 63 participants (76.19%) met the learner criteria on day 1, increasing to 58 out of 63 participants (92.06%) on day 2- Similarly, in the HC group, 34 out of 46 participants (73.91%) reached the learner threshold on day 1, and 44 out of 46 participants (95.65%) achieved it on day 2.These results confirm robust instrumental learning across both groups, with no group-related differences that could confound subsequent PIT effects.

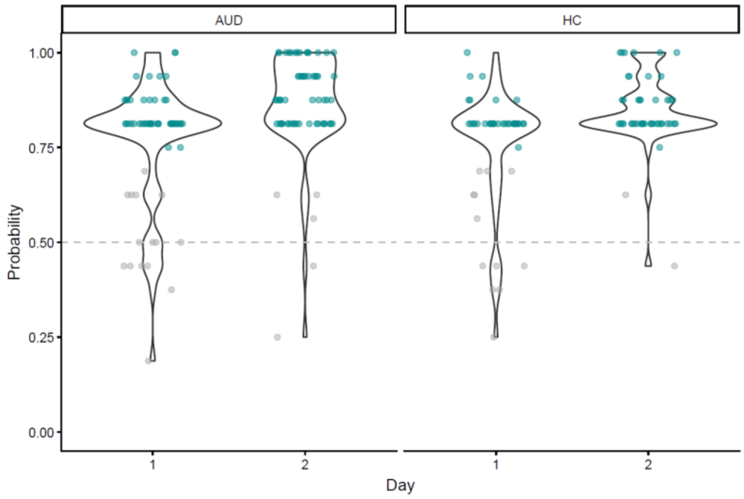


**Figure S4. Instrumental Learning (Mean Correct During Last 16 Trials).** Learner (cyan green), Non-Learner (grey)

### Pavlovian Learning

#### Implicit contingency awareness

Implicit contingency awareness was assessed via the percentage of correct choices of higher-valued CS during the Query trials (PIT task part 4). To evaluate differences in contingency awareness, measured as the percentage of higher-valued CS choices, we conducted Mann–Whitney U tests for each factor: group, intervention, and day.

Overall, the mean contingency awareness was 84.7% (SD = 0.24, Median = 1), with the AUD group obtaining 84.8% correct choices and the HC group 84.6%. The mean correct choice rate was 83.4% on day one and 86.0% on day two. The Mann–Whitney U test indicated no significant differences between groups (W = 5684, p = .655), interventions (W = 5857, p = .553), or days (W = 5614, p = .993).

### S12 Motivational PIT: Random-Effect Structure

**Random Effects Structure**

Linear mixed-effects models were fitted using the lme4 package in R (Bates et al., 2015b). Random-effects structures were selected using a stepwise model comparison procedure following recommendations by Bates et al. 2015 and Matuschek et al. 2017 (Bates et al., 2015a, Matuschek et al., 2017). Models were fitted using maximum likelihood estimation and compared using likelihood ratio tests (LRTs), Akaike's Information Criterion (AIC) and Bayesian Information Criterion (BIC).

The baseline model included a random intercept for day nested within subject: (1 | day:subject. Subsequently, subject random slopes for Pavlovian stimulus and instrumental response were added: (1 + PavStim + InstrResp | subject). This improved model fit relative to the baseline model. Adding a random intercept for day nested within subject further improved fit. To account for item-specific variability additional random intercepts for Pavlovian fractal (Pav_Fractal) and instrumental stimulus (Shell_Leaf) were tested. Inclusion of Pavlovian fractal significantly improved model fit relative to the model containing subject slopes and day. Inclusion of instrumental stimulus yielded a substantial improvement in fit. Finally, adding Pavlovian fractal to the model already containing instrumental stimulus further improved fit, while adding instrumental stimulus to the model already containing Pavlovian fractal also significantly improved fit. The final model (model_full) exhibited the lowest information criteria among all candidate models (AIC = 407791, BIC = 408001) and did not exhibit singularity, indicating an appropriate balance between model complexity and data support. The final random-effects structure was: (1 + PavStim + InstrResp | subject) + (1 | day:subject) + (1 | Pav_Fractal) + (1 | Shell_Leaf).

**Table S8. Model Comparisons Random Effect Structure**

| Model | Random effects | npar | AIC | BIC | $\Delta\chi^2$ | df | p |
|---|---|---|---|---|---|---|---|
| model_base | (1 \| day:subj) | 18 | 411925 | 412070 | – | – | – |
| model_subj | + (1 + PavStim + InstrResp \| subj) | 23 | 409047 | 409232 | 2888.0 | 5 | < .001 |
| model_day | + (1 \| day:subj) | 24 | 408226 | 408419 | 824.0 | 1 | < .001 |
| model_pf | + (1 \| Pav_Fractal) | 25 | 408222 | 408423 | 6.15 | 1 | .013 |
| model_sl | + (1 \| Shell_Leaf) | 25 | 407796 | 407997 | 432.32 | 1 | < .001 |
| model_full | + (1 \| Pav_Fractal) + (1 \| Shell_Leaf) | 26 | 407791 | 408001 | 6.31 / 432.49 | 1 | .012 / < .001 |

### S13 Motivational PIT: Detailed Results and Exploratory Cortisol Analysis

#### Table S9: Results Motivational PIT

| term | estimate | std.error | statistic | df | p.value |
|---|---|---|---|---|---|
| (Intercept) | -260.46 | 120.76 | -2.16 | 72.79 | 0.034 |
| Pavlovian Stimuli | 211.70 | 40.56 | 5.22 | 107.26 | 0.000 |
| Instrumental Response (Go/Push) | 1,945.07 | 131.56 | 14.79 | 146.86 | 0.000 |
| intervention (TSST) | -82.06 | 85.34 | -0.96 | 253.25 | 0.337 |
| group (AUD) | 104.98 | 194.80 | 0.54 | 108.35 | 0.591 |
| Pavlovian Stimuli x Instrumental Response (Go/Push) | 7.80 | 24.85 | 0.31 | 22,768.52 | 0.754 |
| Pavlovian Stimuli x intervention (TSST) | 53.36 | 25.11 | 2.13 | 21,647.15 | 0.034 |
| Instrumental Response (Go/Push) x intervention (TSST) | -114.58 | 107.43 | -1.07 | 22,772.32 | 0.286 |
| Pavlovian Stimuli x group (AUD) | -127.36 | 81.16 | -1.57 | 107.49 | 0.120 |
| Instrumental Response (Go/Push) x group (AUD) | -183.98 | 263.10 | -0.70 | 146.83 | 0.485 |
| intervention (TSST) x group (AUD) | -76.06 | 170.71 | -0.45 | 252.80 | 0.656 |
| Pavlovian Stimuli x Instrumental Response (Go/Push) x intervention (TSST) | -10.08 | 49.70 | -0.20 | 22,768.53 | 0.839 |
| Pavlovian Stimuli x Instrumental Response (Go/Push) x group (AUD) | 35.20 | 49.70 | 0.71 | 22,768.61 | 0.479 |
| Pavlovian Stimuli x intervention (TSST) x group (AUD) | 6.38 | 50.24 | 0.13 | 21,639.66 | 0.899 |
| Instrumental Response (Go/Push) x intervention (TSST) x group (AUD) | 172.39 | 215.02 | 0.80 | 22,774.15 | 0.423 |
| Pavlovian Stimuli x Instrumental Response (Go/Push) x intervention (TSST) x group (AUD) | -22.48 | 99.40 | -0.23 | 22,768.55 | 0.821 |
| sd__(Intercept) | 481.37 | | | | |
| sd__(Intercept) | 903.91 | | | | |
| cor__(Intercept).Pavlovian Stimuli | -0.97 | | | | |
| cor__(Intercept).Instrumental Response | 0.44 | | | | |
| sd__Pavlovian Stimuli | 399.18 | | | | |
| cor__Pavlovian Stimuli.Instrumental Response | -0.42 | | | | |
| sd__Instrumental Response (Go/Push) | 1,242.64 | | | | |
| sd__(Intercept) | 250.78 | | | | |
| sd__(Intercept) | 53.85 | | | | |
| sd__Observation | 1,524.21 | | | | |

**Exploratory Analysis of Cortisol Responder**

To analyse how cortisol responder status modified the PIT effect, the statistical model and its random effect structure remained the same as the one built for monetary trials with the only differences that the fixed effect group (AUD, HC) was replaced by the fixed effect responder (cortisol responder, cortisol non-responder).

**Table S10. Motivational PIT Cortisol Responder vs. Non-Responder**

| term | estimate | std.error | statistic | df | p.value |
|---|---|---|---|---|---|
| (Intercept) | 186.62 | 85.32 | 2.19 | 23.96 | 0.039 |
| Pavlovian Stimuli | 219.69 | 43.10 | 5.10 | 102.32 | 0.000 |
| Instrumental Response (Go/Push) | 1,919.17 | 127.56 | 15.05 | 103.03 | 0.000 |
| intervention (TSST) | 10.07 | 73.03 | 0.14 | 102.62 | 0.891 |
| cortisol responder | 136.34 | 89.41 | 1.52 | 103.88 | 0.130 |
| Pavlovian Stimuli x Instrumental Response (Go/Push) | 19.59 | 26.35 | 0.74 | 21,785.69 | 0.457 |
| Pavlovian Stimuli x intervention (TSST) | 52.63 | 26.54 | 1.98 | 21,350.23 | 0.047 |
| Instrumental Response (Go/Push) x intervention (TSST) | -42.59 | 43.28 | -0.98 | 21,803.10 | 0.325 |
| Pavlovian Stimuli x cortisol responder | 139.43 | 86.19 | 1.62 | 102.26 | 0.109 |
| Instrumental Response (Go/Push) x cortisol responder | -517.61 | 255.09 | -2.03 | 102.98 | 0.045 |
| intervention (TSST) x cortisol responder | 24.72 | 146.54 | 0.17 | 103.08 | 0.866 |
| Pavlovian Stimuli x Instrumental Response (Go/Push) x intervention (TSST) | 6.76 | 52.69 | 0.13 | 21,785.90 | 0.898 |
| Pavlovian Stimuli x Instrumental Response (Go/Push) x cortisol responder | 39.59 | 52.69 | 0.75 | 21,785.68 | 0.452 |
| Pavlovian Stimuli x intervention (TSST) x cortisol responder | -122.01 | 53.35 | -2.29 | 19,404.41 | 0.022 |
| Instrumental Response (Go/Push) x intervention (TSST) x cortisol responder | 609.54 | 86.77 | 7.02 | 21,806.18 | 0.000 |
| Pavlovian Stimuli x Instrumental Response (Go/Push) x intervention (TSST) x cortisol responder | 36.76 | 105.39 | 0.35 | 21,785.93 | 0.727 |
| sd__(Intercept) | 482.42 | | | | |
| sd__(Intercept) | 241.89 | | | | |
| cor__(Intercept).Pavlovian Stimuli | -0.27 | | | | |

| term | estimate | std.error | statistic | df | p.value |
|---|---|---|---|---|---|
| cor__(Intercept).Instrumental Response (Go/Push) | 0.27 | | | | |
| sd__Pavlovian Stimuli | 401.36 | | | | |
| cor__Pavlovian Stimuli.Instrumental Response (Go/Push) | -0.40 | | | | |
| sd__Instrumental Response (Go/Push) | 1,230.73 | | | | |
| sd__(Intercept) | 252.41 | | | | |
| sd__(Intercept) | 66.01 | | | | |
| sd__Observation | 1,527.80 | | | | |
